## Supplementary tables for "Estimating the global demand curve for a leishmaniasis vaccine: a generalisable approach based on global burden of disease estimates"

*Supplementary Table 1: Vaccine rollout projection (2030-2040)*

| | Number of people vaccinated between 2030 and 2040 (for CL prevention) | | | Number of people vaccinated between 2030 and 2040 (for VL prevention) | | |
|---|---|---|---|---|---|---|
| **Country** | **0-4 years** | **5-14 years** | **15-29 years** | **0-4 years** | **5-14 years** | **15-29 years** |
| Afghanistan | 2,749,122 | 5,885,515 | 4,328,652 | - | - | - |
| Algeria | 1,957,406 | 5,163,208 | 2,047,082 | - | - | - |
| Bangladesh | - | - | - | 3,258,918 | 8,191,346 | - |
| Brazil | - | - | - | 10,502,595 | 23,450,987 | - |
| China | - | - | - | 23,028,828 | 53,370,973 | - |
| Ethiopia | 1,812,940 | 2,876,160 | 1,900,381 | 1,138,038 | 1,805,454 | - |
| Georgia | - | - | - | 309,726 | 626,678 | - |
| India | 20,186,202 | 40,113,892 | 27,707,496 | 14,573,985 | 32,893,737 | - |
| Israel | 2,173,296 | 4,419,907 | 2,299,880 | - | - | - |
| Kenya | - | - | - | 1,182,557 | 2,186,535 | - |
| Morocco | 1,267,528 | 2,450,203 | 1,160,361 | - | - | - |
| Nepal | - | - | - | 3,293,786 | 8,223,698 | - |
| Nigeria | 1,305,681 | 2,560,543 | 1,149,318 | - | - | - |
| Pakistan | 22,071,512 | 49,465,950 | 29,566,359 | - | - | - |
| Paraguay | - | - | - | 642,108 | 1,436,384 | - |
| Saudi Arabia | 643,958 | 1,558,356 | 922,762 | - | - | - |
| Somalia | - | - | - | 513,476 | 1,071,733 | - |
| South Sudan | - | - | - | 305,306 | 743,962 | - |
| Spain | - | - | - | 3,059,899 | 7,111,607 | - |
| Sudan | 15,699,043 | 20,950,603 | 13,581,912 | 2,028,979 | 3,452,805 | - |
| Syria | 4,732,334 | 11,634,572 | 5,266,336 | - | - | - |
| Tunisia | 981,419 | 2,287,679 | 1,178,702 | - | - | - |
| Turkey | 7,182,351 | 11,545,177 | 5,136,730 | - | - | - |
| Uzbekistan | 3,378,644 | 8,001,227 | 4,350,362 | - | - | - |
| Total | **564,054,859** | | | | | |

Source: Malvolti S, Malhame M, Mantel C, Rutte EA Le, Kaye PM. Human leishmaniasis vaccines: use cases, target population and potential global demand. PLoS Negl Trop Dis. 2021;Forthcoming.

*Supplementary Table 2: Projected Cost-effectiveness Thresholds (CETs) - 2030 - 2040 (2019 USD)*

| | **Cost-effectiveness Threshold (CET)** | | | | | | | | | | |
|---|---|---|---|---|---|---|---|---|---|---|---|
| **Country** | **2030** | **2031** | **2032** | **2033** | **2034** | **2035** | **2036** | **2037** | **2038** | **2039** | **2040** |
| Afghanistan | 105 | 106 | 107 | 108 | 109 | 110 | 111 | 112 | 113 | 114 | 114 |
| Algeria | 6,522 | 6,609 | 6,703 | 6,793 | 6,877 | 6,953 | 7,026 | 7,102 | 7,165 | 7,233 | 7,300 |
| Bangladesh | 289 | 302 | 315 | 329 | 343 | 358 | 373 | 389 | 407 | 424 | 443 |
| Brazil | 8,766 | 8,820 | 8,858 | 8,878 | 8,904 | 8,935 | 8,966 | 9,002 | 9,036 | 9,074 | 9,097 |
| China | 12,386 | 13,069 | 13,774 | 14,525 | 15,304 | 16,117 | 16,961 | 17,835 | 18,739 | 19,701 | 20,706 |
| Ethiopia | 443 | 464 | 485 | 509 | 534 | 560 | 587 | 615 | 645 | 677 | 710 |
| Georgia | 1,285 | 1,309 | 1,333 | 1,361 | 1,390 | 1,418 | 1,447 | 1,477 | 1,504 | 1,537 | 1,571 |
| India | 676 | 707 | 742 | 777 | 813 | 852 | 893 | 936 | 982 | 1,029 | 1,078 |
| Israel | 5,626 | 5,700 | 5,774 | 5,847 | 5,919 | 5,991 | 6,067 | 6,143 | 6,219 | 6,296 | 6,372 |
| Kenya | 843 | 860 | 877 | 893 | 910 | 927 | 943 | 958 | 974 | 990 | 1,006 |
| Morocco | 1,945 | 2,012 | 2,078 | 2,143 | 2,212 | 2,285 | 2,361 | 2,438 | 2,511 | 2,590 | 2,670 |
| Nepal | 419 | 429 | 439 | 450 | 461 | 472 | 484 | 496 | 507 | 520 | 532 |
| Nigeria | 278 | 279 | 281 | 282 | 284 | 285 | 286 | 288 | 289 | 291 | 292 |
| Pakistan | 244 | 251 | 259 | 267 | 275 | 284 | 293 | 301 | 310 | 320 | 329 |
| Paraguay | 7,801 | 8,020 | 8,248 | 8,490 | 8,722 | 8,979 | 9,229 | 9,491 | 9,740 | 10,009 | 10,294 |
| Saudi Arabia | 2,554 | 2,546 | 2,538 | 2,526 | 2,514 | 2,499 | 2,488 | 2,477 | 2,464 | 2,451 | 2,437 |
| Somalia | 24 | 25 | 25 | 25 | 25 | 25 | 25 | 25 | 25 | 25 | 25 |
| South Sudan | 81 | 81 | 82 | 82 | 82 | 82 | 83 | 83 | 83 | 83 | 83 |
| Spain | 3,604 | 3,609 | 3,613 | 3,618 | 3,625 | 3,629 | 3,634 | 3,639 | 3,644 | 3,649 | 3,653 |
| Sudan | 484 | 490 | 497 | 503 | 509 | 515 | 520 | 525 | 530 | 535 | 540 |
| Syria | 231 | 232 | 233 | 232 | 231 | 229 | 229 | 228 | 227 | 226 | 224 |
| Tunisia | 4,409 | 4,452 | 4,495 | 4,535 | 4,577 | 4,618 | 4,657 | 4,687 | 4,714 | 4,742 | 4,762 |
| Turkey | 17,561 | 18,146 | 18,758 | 19,418 | 20,130 | 20,815 | 21,538 | 22,291 | 23,093 | 23,890 | 24,742 |
| Uzbekistan | 1,953 | 2,012 | 2,070 | 2,131 | 2,193 | 2,258 | 2,317 | 2,384 | 2,453 | 2,522 | 2,591 |

*Supplementary Table 3: Projected GAVI support status in 2030*

| **Country** | **Projected country support status in 2030** |
|---|---|
| Afghanistan | Initial self-financing |
| Algeria | Not eligible |
| Bangladesh | Preparatory transition |
| Brazil | Not eligible |
| China | Not eligible |
| Ethiopia | Preparatory transition |
| Georgia | Not eligible |
| India | Accelerated transition |
| Israel | Not eligible |
| Kenya | Accelerated transition |
| Morocco | Not eligible |
| Nepal | Preparatory transition |
| Nigeria | Accelerated transition |
| Pakistan | Accelerated transition |
| Paraguay | Not eligible |
| Saudi Arabia | Not eligible |
| Somalia | Initial self-financing |
| South Sudan | Initial self-financing |
| Spain | Not eligible |
| Sudan | Not eligible |
| Syria | Preparatory transition |
| Tunisia | Not eligible |
| Turkey | Not eligible |
| Uzbekistan | Not eligible |
